# Persistence of robust humoral immune response in COVID-19 convalescent individuals over 12 months after infection

**DOI:** 10.1101/2021.09.27.21264013

**Authors:** Kei Miyakawa, Sousuke Kubo, Sundararaj Stanleyraj Jeremiah, Hirofumi Go, Yutaro Yamaoka, Norihisa Ohtake, Hideaki Kato, Satoshi Ikeda, Takahiro Mihara, Ikuro Matsuba, Naoko Sanno, Masaaki Miyakawa, Masaharu Shinkai, Tomoyuki Miyazaki, Takashi Ogura, Shuichi Ito, Takeshi Kaneko, Kouji Yamamoto, Atsushi Goto, Akihide Ryo

**Affiliations:** Department of Microbiology, Yokohama City University Graduate School of Medicine, Kanagawa, Japan; Department of Pulmonology, Yokohama City University Graduate School of Medicine, Kanagawa, Japan; Department of Biostatistics, Yokohama City University Graduate School of Medicine, Kanagawa, Japan; Life Science Laboratory, Technology and Development Division, Kanto Chemical Co, Inc., Kanagawa, Japan; Advanced Medical Research Center, Yokohama City University, Yokohama, Japan; Bioscience Division, Research and Development Department, Tosoh Corporation, Tokyo Research Center, Kanagawa, Japan; Infection Prevention and Control Department, Yokohama City University Hospital, Kanagawa, Japan; Department of Respiratory Medicine, Kanagawa Cardiovascular and Respiratory Center, Kanagawa, Japan; Department of Health Data Science, Yokohama City University Graduate School of Data Science, Kanagawa, Japan; Matsuba Medical Clinic, Kanagawa, Japan; Shinagawa Strings Clinic, Tokyo, Japan; Miyakawa Internal Medicine and Pediatrics Clinic, Kanagawa, Japan; Japan Medical Association, Tokyo, Japan; Division of Internal Medicine, Tokyo-Shinagawa Hospital, Tokyo, Japan; Department of Physiology, Yokohama City University Graduate School of Medicine, Kanagawa, Japan; Department of Pediatrics, Yokohama City University Graduate School of Medicine, Kanagawa, Japan

**Author notes:** **Corresponding authors:** Kei Miyakawa, Atsushi Goto, Akihide Ryo **[lead contact]**. These authors contributed equally to this work (Kei Miyakawa, Sousuke Kubo, Sundararaj Stanleyraj Jeremiah).

## Abstract

SARS-CoV-2 infection elicits varying degrees of protective immunity conferred by neutralizing antibodies (nAbs). Here we report the persistence of nAb responses over 12 months after infection despite its decreasing trend noticed from 6 months. The study included sera from 358 individuals who had been infected with SARS-CoV-2 between January and May 2020. Samples were collected at 6 and 12 months after onset. The titers of IgG to the viral nucleocapsid protein (NP) and receptor-binding domain of the spike protein (RBD) were measured by CLEIA. The nAb titer was determined using lentivirus-based pseudovirus or authentic virus. Antibody titers of NP-IgG, RBD-IgG, and nAbs were higher in severe and moderate cases than in mild cases at 12 months after onset. While the nAb levels were likely to confer adequate protection against wild-type viral infection, the neutralization activity to recently circulating variants in some of the mild cases (∼30%) was undermined, implying the susceptibility of reinfection to the variants of concerns (VOCs). COVID-19 convalescent individuals have robust humoral immunity even at 12 months after infection albeit that the medical history and background of patients could affect the function and dynamics of antibody response to the VOCs.

## Introduction

Severe acute respiratory syndrome coronavirus 2 (SARS-CoV-2), the causative agent of the coronavirus disease 2019 (COVID-19) pandemic, has infected nearly 200 million people worldwide, causing over 4 million deaths, as of the end of July 2021 (https://coronavirus.jhu.edu/). Vaccines are the only limiting modality [1]. Because of to the inability of equitable distribution and rapid mass vaccination, the virus is spreading almost unchecked and has led to the emergence of variant mutants [2-4]. Some of the variants pose significant epidemiological problems, and the World Health Organization (WHO) has defined these strains as variants of concern (VOCs) or variants of interest (VOIs). Since both naturally acquired post-infection immunity and artificially acquired vaccine-mediated immunity produce neutralizing antibodies (nAbs), which are helpful in limiting the future course of the pandemic, it is important to know about the long-term persistence and assess the neutralizing activity of infection and vaccine-derived immunity against VOCs and VOIs. Several studies have investigated the longevity of post-infection nAb titers, but only a few long-term follow-up studies have been conducted, especially against the VOCs and VOIs, one year after infection [5-7].

In this study, we collected blood samples from 358 patients diagnosed with COVID-19 in Japan at 6 and 12 months after the onset of the disease and performed a comprehensive analysis of humoral immunity. We examined levels of immunoreactive antibodies targeting SP-RBD and NP, as well as nAbs against multiple VOCs and VOIs. We also investigated host factors that influence on the persistence of antibody response.

## Results

### Study cohort

The study included sera from 358 volunteers who had been infected with SARS-CoV-2 between January and May 2020, confirmed either by reverse transcription polymerase chain reaction (RT-PCR), reverse transcription loop-mediated isothermal amplification (RT-LAMP), or antigen testing. The mean age of the participants was 50.1 ± 13.2 years with a male to female ratio of 54.5% to 45.5%. Demographic data and acute-phase symptoms of the cohort are shown in **Supplementary Table S1**. According to the WHO’s disease severity definitions [8], the participants were assigned as asymptomatic/mild (74% [264/358]), moderate (19% [69/358]), or severe (7% [25/358]) (**Fig. 1A**).

**Figure 1.**
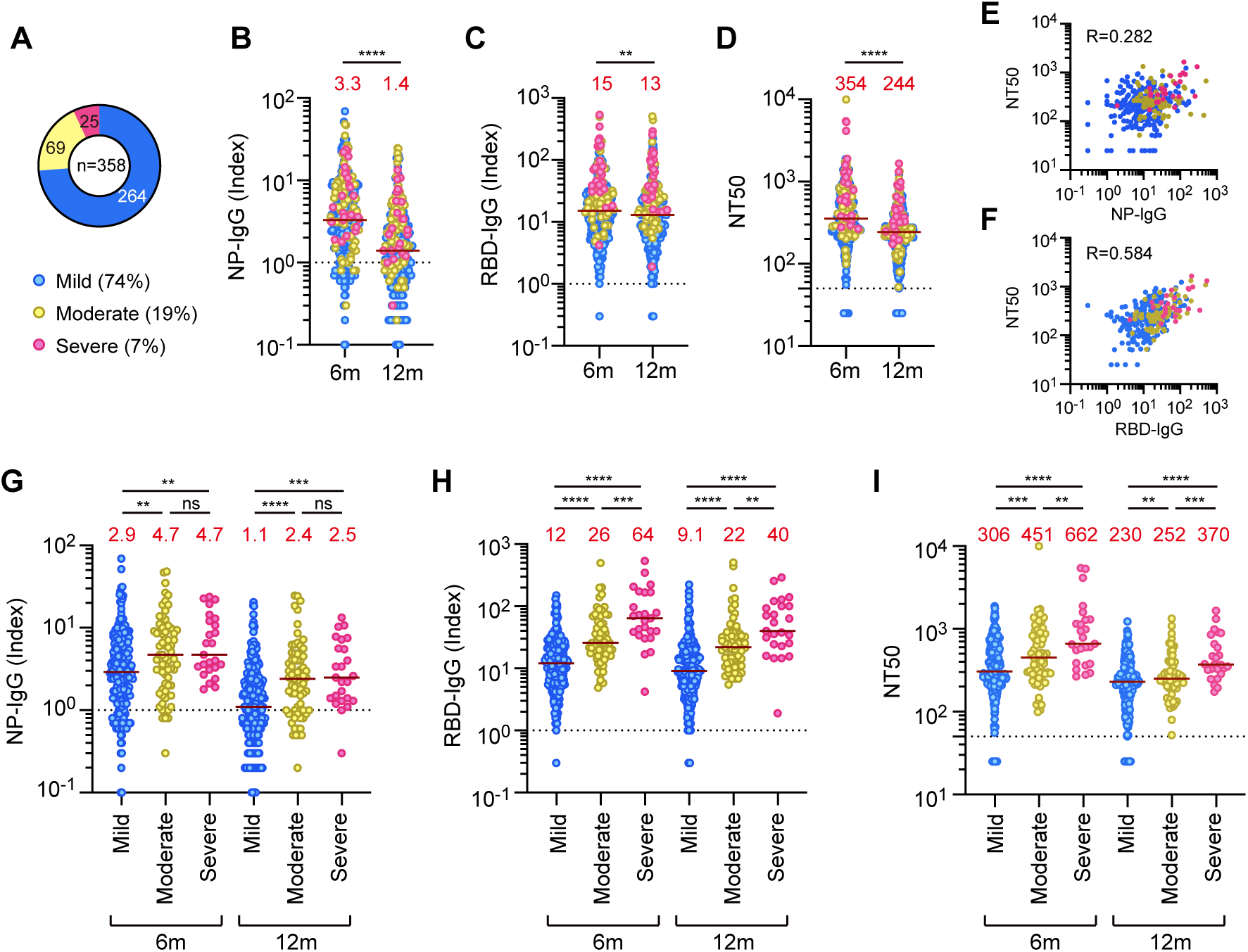
Severity of COVID-19 and persistence of SARS-CoV-2 antibody. **(A)** Severity of acute phase in the study participants (total n = 358). **(B-D)** Amounts of SARS-CoV-2 NP-IgG (B), RBD-IgG (C), and neutralizing activity (NT50) (D) in COVID-19 convalescent sera at 6 and 12 months after onset (total n = 358). Dotted lines indicate threshold values for each antibody. Blue, yellow, and pink indicate mild, moderate, and severe patients, respectively. Red letters on the graph indicate the median value of each antibody titer. \*\**P* < 0.01; \*\*\*\**P* < 0.0001; ns, not significant, two-tailed paired t-test. **(E, F)** Correlation between NT50 and NP-IgG (E) or RBD-IgG (F) in COVID-19 convalescent sera at 12 months after onset. **(G-I)** Relationship between COVID-19 severity and the amounts of NP-IgG (G), RBD-IgG (H), or NT50 (I) in COVID-19 convalescent sera (total n = 358). \**P* < 0.05; \*\**P* < 0.01; \*\*\**P* < 0.001; \*\*\*\**P* < 0.0001; ns, not significant, two-tailed unpaired t-test.

### Severity of COVID-19 and persistence of SARS-CoV-2 antibody

The presence of serum IgG against the viral nucleocapsid protein (NP) or receptor-binding domain (RBD) of the viral spike protein (SP) was quantitatively examined using the Tosoh AIA-CL chemiluminescence enzyme immunoassay (CLEIA) [9]. Median NP-IgG titer was 3.3 at 6 months and 1.4 at 12 months (**Fig. 1B)**, and that of RBD-IgG was 15.2 at 6 months and 13.0 at 12 months (**Fig. 1C**). Both titers decreased significantly between 6 and 12 months, but the rate of decrease was higher for NP-IgG than for RBD-IgG. Next, we examined the neutralizing titer (NT50) values of these sera using the well-established HIV-derived pseudovirus neutralization assay to assess the presence of nAbs [10, 11]. Interestingly, we observed that the nAbs were maintained after 12 months from their levels at 6 months (**Fig. 1D**), which was similar to the trend observed for RBD-IgG (**Fig. 1C**). NT50 values showed a definite correlation with RBD-IgG titers, but not with NP-IgG titers (**Fig. 1E, F**). When antibody titers of NP-IgG, RBD-IgG, and nAbs were classified according to symptoms, all of them were higher in severe and moderate cases than in mild cases at both the tested time periods (**Fig. 1G, H, I**).

### Host factors versus persistence of SARS-CoV-2 antibodies

The study participants had a wide variation in age, ranging from the 20s to the 70s, and increasing rates of moderate and severe cases were observed starting from the 40s and above (**Fig. 2A**). In the age-stratified cohort, the titers of NP-IgG, RBD-IgG, and NT50 after 12 months were higher in patients aged > 50 years (**Fig. 2B**). A decrease in nAbs over time was observed in all strata.

**Figure 2.**
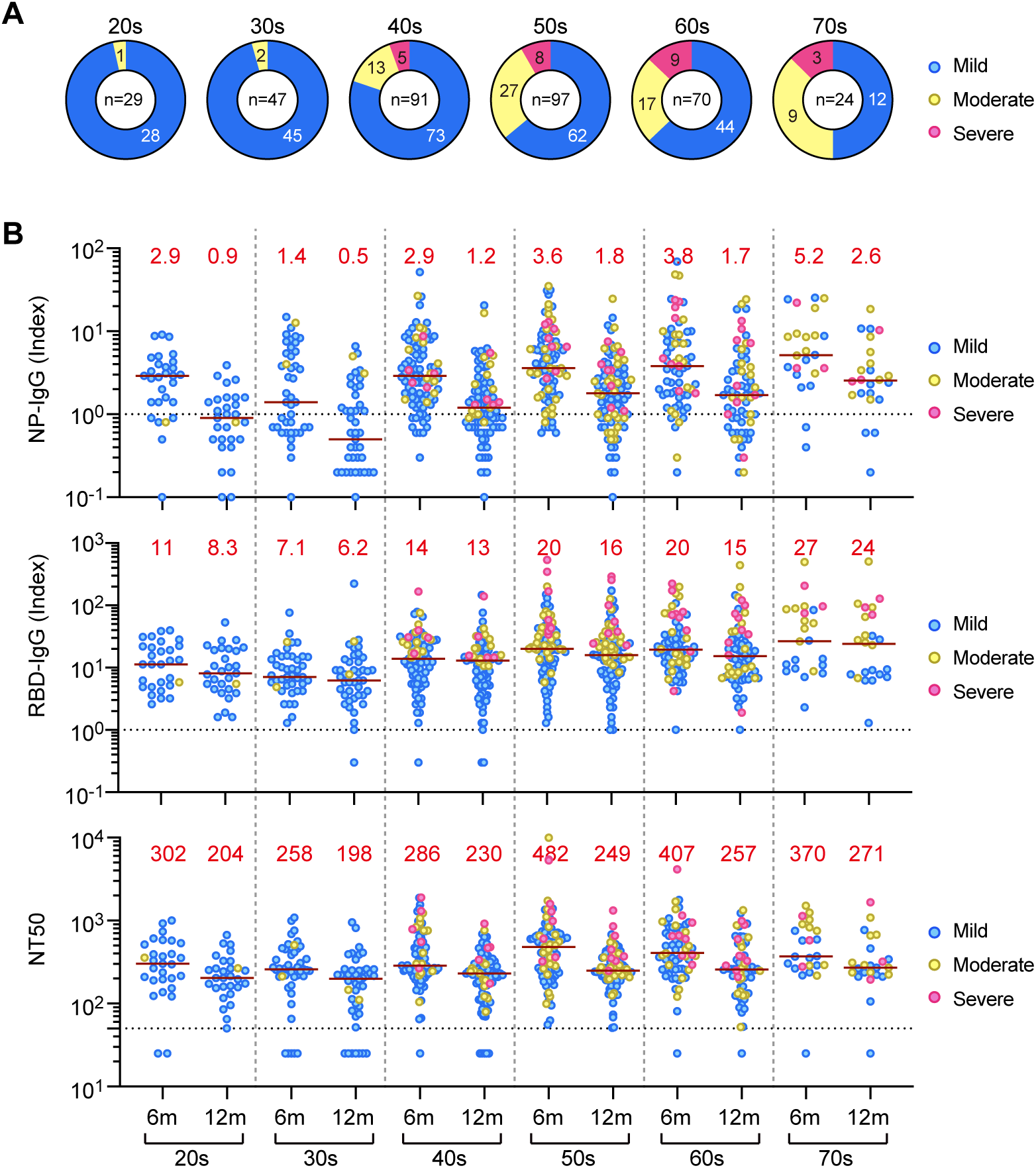
Onset age of COVID-19 and persistence of SARS-CoV-2 antibody. **(A)** Age and acute severity of the participants in this study (total n = 358). **(B-D)** Amounts of NP-IgG (B), RBD-IgG (C), and NT50 (D) by age groups in COVID-19 convalescent sera at 6 and 12 months after onset. Blue, yellow, and pink indicate mild, moderate, and severe patients, respectively. Red letters on the graph indicate the median value of each antibody titer.

Since COVID-19 is known to be more severe in males [12], we examined the antibody titers between the sexes. We observed that males had a higher frequency of severe disease, but no significant difference was observed in the reduction of nAb titers (**Supplementary Fig. S1A**). Disease of increased severity occurred more commonly in those with a higher body mass index (BMI) and in smokers, but there were no significant differences in the 12 months nAb titers in both cohorts (**Supplementary Fig. S1B, C**). We conducted a multiple regression analysis for examining the association between amount of NT50 at 12 months after onset and host factors, with ordinary log-transformed NT50 as the objective variable and age, sex, BMI, and smoking as explanatory variables, as well as severity of illness during the disease. The results of the analysis showed that the severity of the acute phase and the age of onset were statistically significantly associated with amount of NT50 after 12 months (**Supplementary Table S2**).

### Temporal changes in neutralizing titer in individual subjects

Next, we classified the 358 subjects into four groups based on their nAb titer transition between 6 and 12 months, on the similar lines of Chia et al. [13]: (Group 1 Significant decrease) those whose nAb titers decreased by > 50% (Group 2 Gradual decrease) those whose titers decreased between 20% and 50%, (Group 3 Persistent) those whose titers remained between 20% increase or decrease, (Group 4 Increase) and those whose titers increased by > 20% (**Fig. 3A**). Thirty-three of the 358 (37%) subjects were in Group 1, 24% (85 of 358) were in Group 2, 23% (84 of 358) were in Group 3, and Group 4 included 16% (56 of 358). A major proportion of the subjects (61%) showed a reduction in nAb titers (Groups 1 and 2) at 12 months (**Fig. 3B**). Despite the reduction, effective neutralizing activity (NT50 > 50) was observed in most patients (344 out of 358; 96%) after 12 months, with only a very few falling below the detection limit. When classified according to the symptoms presented during the acute period, most of the patients with severe disease belonged to Group 1, while those of mild cases were spread among all groups (**Fig. 3C**). These results suggest that people presenting with severe disease form higher levels of nAbs, which also decrease at a faster rate. However, despite this decrease, the nAb titer may continue to be maintained at low functional levels.

**Figure 3.**
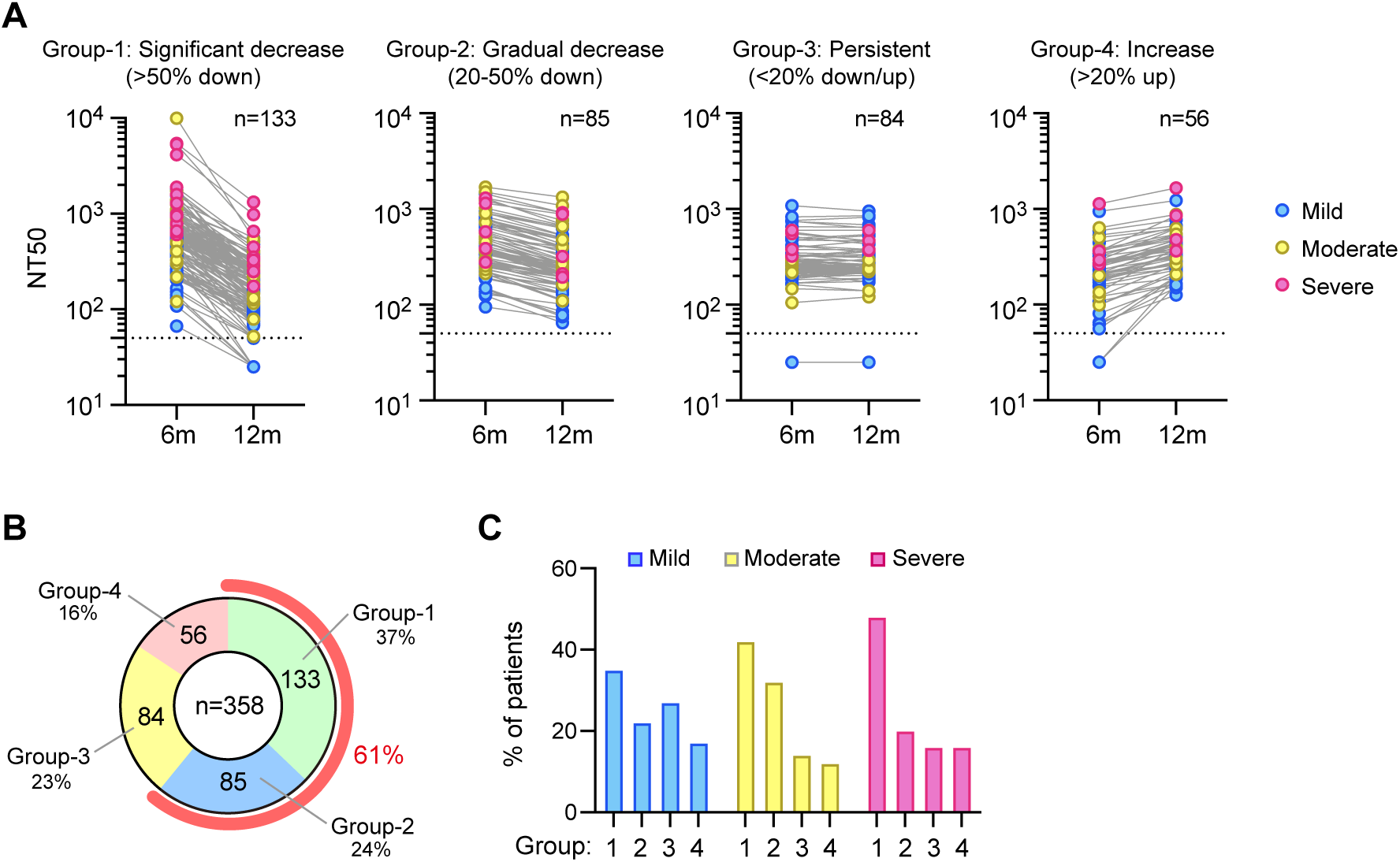
Temporal changes in neutralizing titer in the same patient. **(A)** Classification of 358 patients based on neutralizing activity (NT50). Significantly decreased (Group 1), slowly decreased (Group 2), maintained (Group 3), and increased (Group 4) between 6 and 12 months of onset. Blue, yellow, and pink indicate mild, moderate, and severe patients, respectively. **(B)** Percentage of each group in all subjects. **(C)** Percentage of each group in indicated COVID-19 severity.

### Neutralizing activity against SARS-CoV-2 variants in convalescent sera

Next, we investigated whether nAbs of convalescents produced during past infections (possibly by the initial viral strains A or B.1) had the ability to neutralize the VOCs/VOIs of SARS-CoV-2, namely alpha (B.1.1.7), beta (B.1.351), gamma (P.1), delta (B.1.617.2), and kappa (B.1.617.1) strains, which evolved later. These strains have multiple mutations throughout the length of the spike gene, of which the E484K/Q mutation is possessed by the beta, gamma, and kappa strains and causes immune evasion [14, 15]. The delta strain does not have an E484 mutation, but other mutations such as L452R and T478K accumulated in the RBD may confer this strain with the potential for immune evasion [16]. Using 20 randomly selected paired sera, we observed a significant decrease in neutralizing titer at 12 months against the beta, gamma, and kappa strains, which have an E484K/Q mutation, while the sera retained neutralizing efficacy for the alpha and the currently rampant delta strain (**Fig. 4A**). Notably, convalescents of severe cases had high neutralizing activity against all variants (**Fig. 4A**). We also found that the RBD-IgG titer against the beta and gamma strains was also reduced compared to wild-type and alpha strains (**Fig. 4B**), and NT50 correlated well with the RBD-IgG titer of each VOC (**Supplementary Fig. S2**). These results together suggest that accumulated mutations favor the beta and gamma strains to escape humoral immunity by weakening the binding ability of nAbs. Although Moriyama et al. [17] recently showed that the neutralization potency (NT50 divided by RBD-IgG) of the antibody increases over time as an indicator of the maturation of humoral immunity, we did not observe a robust increase in the neutralizing potency at 6 and 12 months (**Supplementary Fig. S3**). We next examined the positivity rate of nAbs using the rapid qualitative neutralizing assay [18, 19], and found that some convalescents of mild cases (∼30%) lost the nAbs to the VOCs/VOIs with a greater proportion (**Fig. 4C**). Further, convalescents of moderate or severe cases showed a nAb positivity rate of more than 90% for all tested variants even 12 months post-infection (**Fig. 4C**). These results indicate that VOCs/VOIs are particularly capable of evading humoral immunity acquired from earlier infection, but convalescent sera of moderate or severe cases have a higher ability to neutralize these strains compared to those of mild cases.

**Figure 4.**
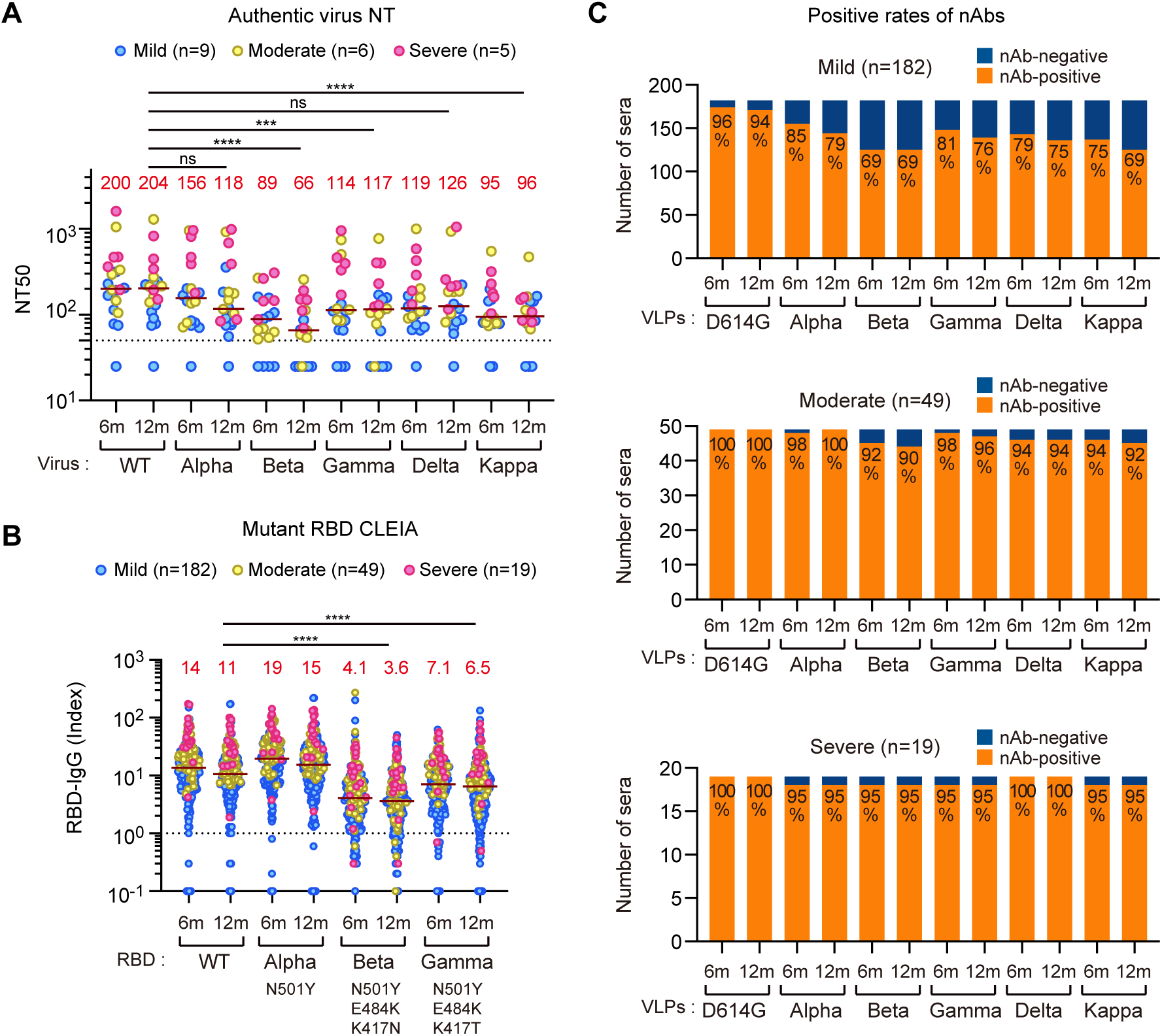
Neutralizing activity against SARS-CoV-2 variants in convalescent sera. **(A)** Neutralizing activity (NT50) against wild-type (WT) SARS-CoV-2 and its variant strains (alpha, beta, gamma, delta, and kappa) in COVID-19 convalescent sera at 6 and 12 months after onset, as revealed by the authentic virus neutralizing test (total n = 20). Red letters on the graph indicate the median value of each antibody titer. \*\**P* < 0.01; ns, not significant, two-tailed paired t-test. **(B)** Amounts of IgG against mutated RBD antigen, as revealed by CLEIA (total n = 250). Red letters on the graph indicate the median value of each antibody titer. \*\*\*\**P* < 0.0001; ns, not significant, two-tailed paired t-test. **(C)** Positive rates of neutralizing antibodies against indicated variants, as revealed by qualitative neutralizing test (total n = 250, see Material and Methods).

### Comparison of neutralizing antibody titers between convalescent and vaccinee sera

We recently reported serological analyses of COVID-19 vaccinees against mutant strains [19]. The nAb titers in these vaccine study cohorts were quantitatively measured and compared to those of COVID-19 convalescents. Previously uninfected vaccinees had a median nAb titer of 76 after the first dose of mRNA vaccine (BNT162b2), which is much lower than that of convalescents. The second dose of the vaccine resulted in a median nAb titer of 841, which was higher than that of convalescents (**Supplementary Fig. S4**). In contrast, previously infected subjects with a median nAb titer of 216 prior to vaccine administration showed a dramatic increase to 4678 after only one dose of mRNA vaccine. Interestingly, previously infected individuals did not experience a further increase in the nAb titer after the second dose of the vaccine. These results indicate that to obtain the equivalent nAb titer as those of convalescents, two doses of vaccination are necessary for previously uninfected persons, and a single dose would suffice for those previously infected.

## Discussion

In this study, we examined the persistence of serum nAbs and their effectiveness in neutralizing VOCs and VOIs at 12 months in 358 naturally infected individuals. Early reports have indicated that nAbs acquired upon infection could disappear within the following 3 months [6]; however, a series of recent reports indicate that they are maintained for longer durations [20-24]. We reveal that nAbs persist at functionally effective titers at one year despite showing a declining trend in most study participants (61%). Our report supports a recent report that infection immunity is maintained for at least one year following natural infection [24], and it further suggests the possibility of nAbs persisting longer. In a cohort of 164 patients, Chia et al. demonstrated that over 65% of subjects showed either a negative or a declining trend in nAbs at 6 months. We observed a similar trend, but at 12 months, which possibly indicates the better detection efficacy of the CLEIA used in our study than the methods employed by Chia et al. [13].

Several reports have suggested that overall antibody levels, including the nAb titer, correlate with the severity of infection [24, 25]. Severe patients have a much higher viral load, which may elicit a more robust humoral response than patients with mild or asymptomatic illness [26]. Consistent with these reports, we observed in our cohort that the individuals who recovered from severe disease possessed higher levels of all types of antibodies tested. These patients had high nAb titers and showed higher neutralizing capacity against VOCs, including the beta strain, notorious for immune escape, and the more rampant delta strain. In contrast, those recovered from mild illness not only had lower nAb levels, but also showed reduced protection against the VOCs. This may be because of nAbs having reduced binding activity against RBD of VOCs.

The duration and intensity of humoral immunity after SARS-CoV-2 infection has been shown to vary based on the severity of the clinical presentation [27]. Some host factors, such as sex, obesity, and smoking, have been reported to influence disease severity [12]. However, the effect of these host factors on the persistence of nAbs at 12 months after infection remains unknown. We reveal in this study that disease severity in the acute phase and the age of the patients correlate significantly with the magnitude of neutralization activity after 12 months. Muller et al. reported that older people have a weaker humoral response to vaccines characterized by lower titers of nAbs in the elderly cohort than in the younger cohort [28]. In contrast, our findings suggest that in natural infection, the elderly possess more nAbs, probably because the elderly more often have serious diseases, which results in the production of more nAbs, which is not the scenario with a fixed dose of antigenic exposure in vaccination. We also found that to obtain the equivalent titer of nAbs as that of the convalescents, two doses of vaccination are necessary for previously uninfected persons, while a single dose could achieve the same in previously infected individuals, corroborating previous suggestions [29].

While the factors responsible for the maintenance of neutralizing activity remain unclear, recent reports suggest that B-cell clones expressing broad and potent nAbs are selectively retained in the repertoire over time [11, 30]. Moriyama et al. suggested that the neutralizing potency (NT50 divided by RBD-IgG) of the antibody increases over time [17]. Although this seems to be an attractive indicator for the maturation of humoral immunity in convalescents, we did not observe a robust increase in the neutralizing potency between 6 and 12 months. This finding suggests that selection of B-cell clones expressing potent nAb may occur immediately after the acute phase, and selection between 6 and 12 months after onset may be rare [24]. Alternatively, the initial high-potency antibodies produced may continue to be produced, especially in those with severe disease.

## Methods

### Experimental design and Ethics statement

Patients with COVID-19 were recruited in the manner described earlier [21]. Briefly, we used our institutional website (not operational at present), social network services, and general mass media to recruit recovering patients. Eligible participants were aged ≥ 20 years at study entry time, resided in Japan, and had a positive result in either RT-PCR, RT-LAMP, or antigen tests for SARS-CoV-2. For all participants, physicians at cooperating outpatient clinics confirmed the diagnosis of COVID-19 based on information provided by their hospitals, clinics, or public health centers. The inclusion and exclusion criteria are available at the University Hospital Medical Information Network-Clinical Trials Registry (UMIN-CTR), where this study was registered (Number UMIN000041227; UMIN Clinical Trials Registry, 2021). The primary endpoints of this study were NT50 and titers of antibodies against NP and SP antigens at 20–32 weeks (visit 1) and 46– 58 weeks (visit 2) after the first positive SARS-CoV-2 test results. In this report, participants who provided their serum samples from January 25th to March 24th, 2021, at visit 2 were included. Demographic data and acute-phase symptoms of the cohort are shown in **Supplementary Table S1**. All participants provided written informed consent and the study was approved by the Institutional Review Board of Yokohama City University (Reference No. B200600115, B200700023, B210300001).

### Serological testing for SARS-CoV-2 NP-IgG and RBD-IgG

The amounts of SARS-CoV-2 NP-IgG and RBD-IgG in sera were quantified using the automated immunoassay AIA-CL1200 (Tosoh). In this study, the threshold index was set to 1 based on previous validations [9], which has been shown to have high sensitivity and specificity. To detect IgG against variant RBD, the antigen with a single mutation (N501Y) or triple mutations (N501Y, E484K, K417N/T) were expressed in Expi293F cells (Thermo Fisher Scientific), according to the manufacturer’s instructions. The supernatants, including recombinant proteins, were harvested and purified using a Strep-tag purification system (IBA Lifesciences). The conjugation of RBD proteins to micromagnetic beads and their attachment to the AIA-CL instrument have been described previously [9].

### Neutralizing assays

Neutralizing assay using peudovirus and rapid qualitative neutralizing assay were performed as previously described [18, 19, 21]. Authentic SARS-CoV-2 was obtained from the National Institute of Infectious Diseases in Japan and handled at the BSL3 facility. Details of the strains used in this study are listed in **Supplementary Table 3**. For the neutralizing assay, VeroTM cells seeded in 96-well plates were washed and infected with 100 µL of medium containing authentic SARS-CoV-2 (moi = 0.05) and five-fold serially diluted serum (1:50–1:31,250 dilution). Three days after infection, cells were washed and 40 µL of CellTiter-Glo Substrate (Promega) was added. Cell viability was measured using the GloMax Discover System (Promega). The calculation method for NT50 was described above.

### Statistical Analysis

All statistical analysis was performed using Prism v8.3.1 (GraphPad) and R version 4.0.3 (R, Foundation for Statistical Computing). Statistical test for each analysis was described in each legend. Statistical significance was defined as p < 0.05.

## Data Availability

The datasets presented in this article are not readily available because it is difficult to ensure the de-identification of data. However, they can be available from the corresponding authors on reasonable request.

## Data sharing

The datasets presented in this article are not readily available because it is difficult to ensure de-identification of data. However, they can be available from the corresponding authors on reasonable request.

## Author contributions

Conceptualization: KM, HG, AG, AR

Methodology: KM, SK, YY, NO, AR

Research design and sample collection: HK, SIkeda, TMihara, IM, NS, MM, MS, AG

Data Analysis: KM, SK, SSJ, HG, YY, NO, HK, TMihara, TMiyazaki, SIto, TK, KY

Writing—original draft: KM, SSJ

Writing—review & editing: KM, SSJ, AG, AR

## Declaration of Competing Interest

YY is a current employee of Kanto Chemical Co., Inc, and NO is a current employee of Tosoh Corporation. The remaining authors declare that the research was conducted in the absence of any commercial or financial relationships that could be construed as a potential conflict of interest.

## Acknowledgements

We acknowledge all the patients and medical staff involved in this study. Cooperating clinics are listed in the **Supplementary Appendix**. We thank Kazuo Horikawa, Kenji Yoshihara, and Natsumi Takaira for their technical assistance.

## Funding

This research was supported by AMED under grant numbers JP19fk0108110 and JP20he0522001.

## Supplementary information

**Supplementary Figure S1.**
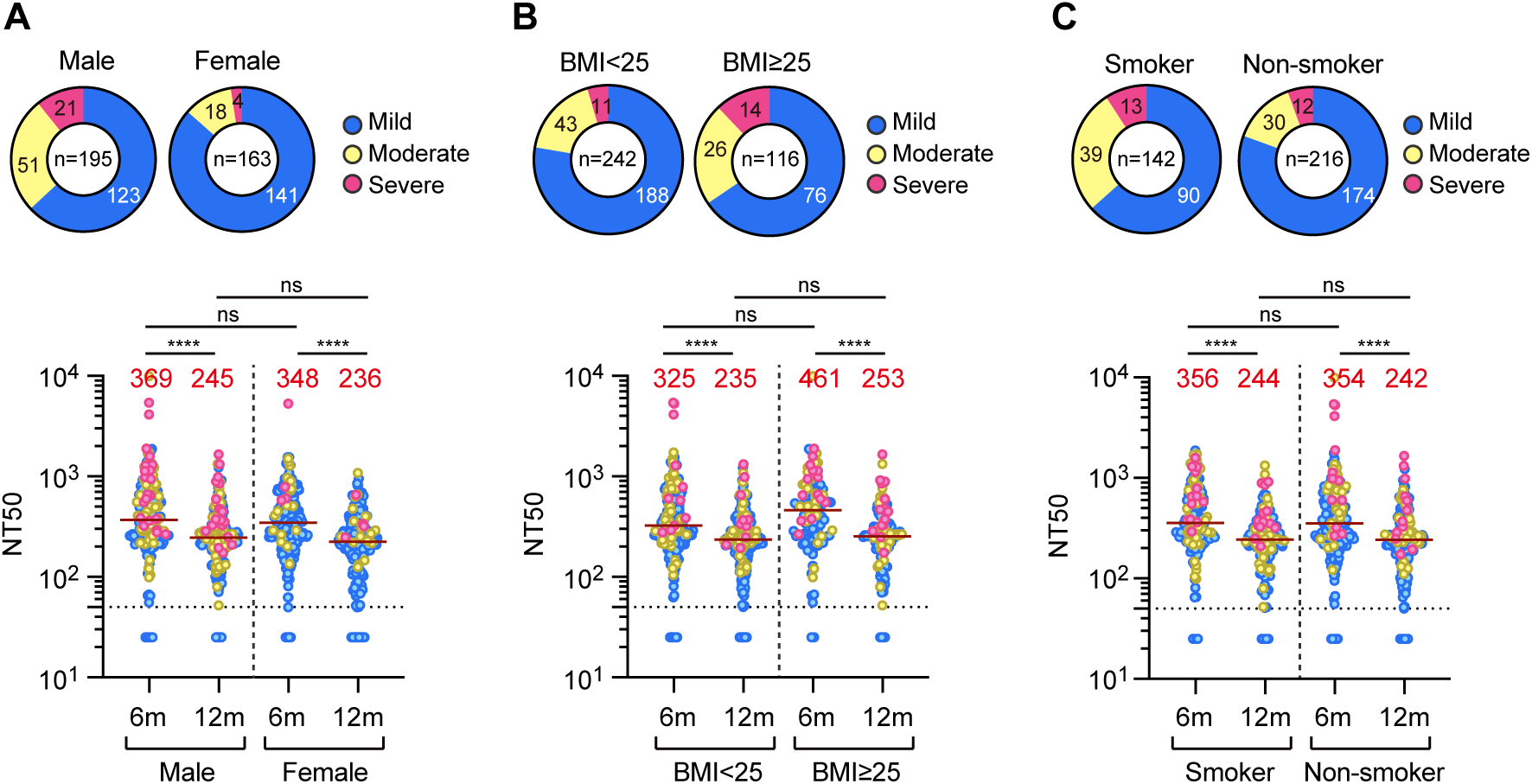
Effect of sex, BMI, and smoking history on maintenance of neutralizing antibodies. (A-C) Amounts of NT50 by groups of sex (A), BMI (B), and smoking history (C) in COVID-19 convalescent sera at 6 and 12 months after onset. Acute severity of the participants in this study was also shown. Blue, yellow, and pink indicate mild, moderate, and severe patients, respectively. Red letters on the graph indicate the median value of each antibody titer. \*\*\*\**P* < 0.0001; ns, not significant, two-tailed unpaired t-test.

**Supplementary Figure S2.**
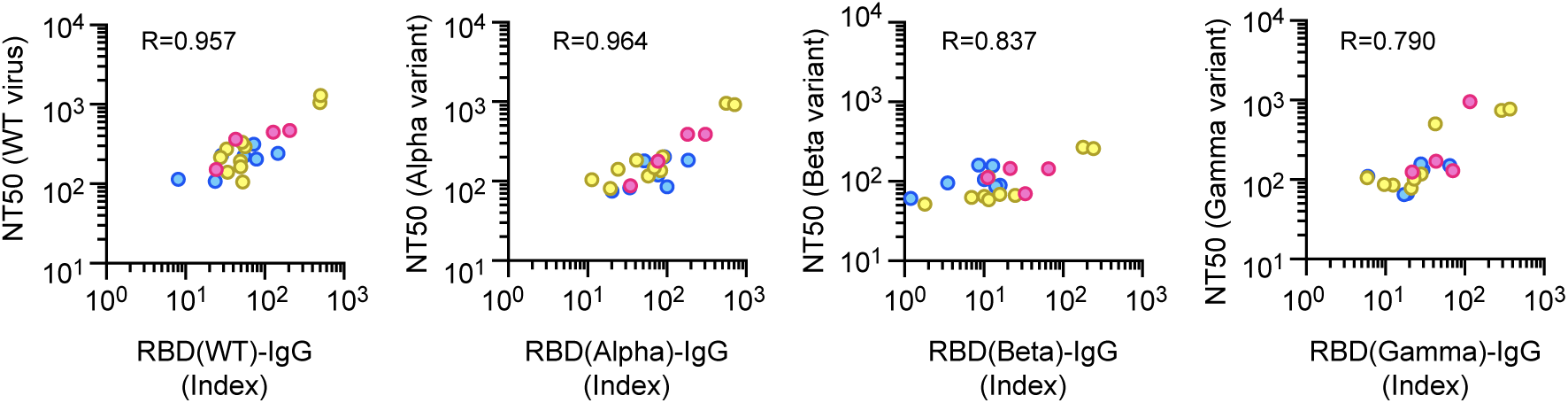
Correlation between NT50 and RBD-IgG against indicated variants. NT50 against VOCs correlated well with the RBD-IgG titer of each VOC.

**Supplementary Figure S3.**
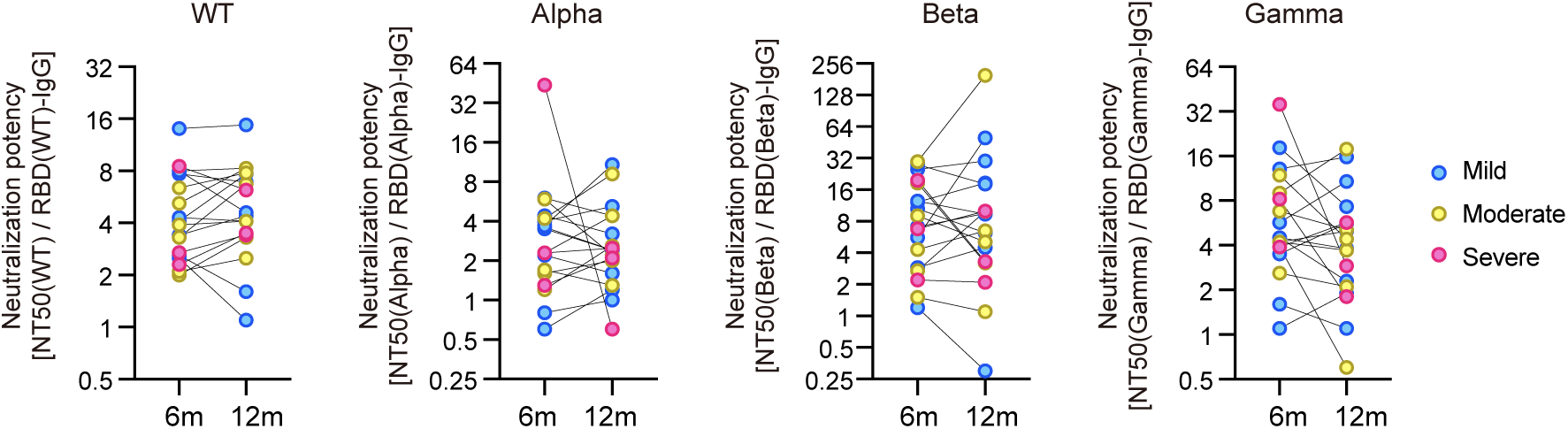
Temporal changes in neutralization potency index in the same patient. Changes in the neutralization potency index (NPI) at 6 and 12 months after onset in the same patient are shown by the indicated variants of SARS-CoV-2. NPI was calculated by dividing each NT50 by the RBD-IgG titer.

**Supplementary Figure S4.**
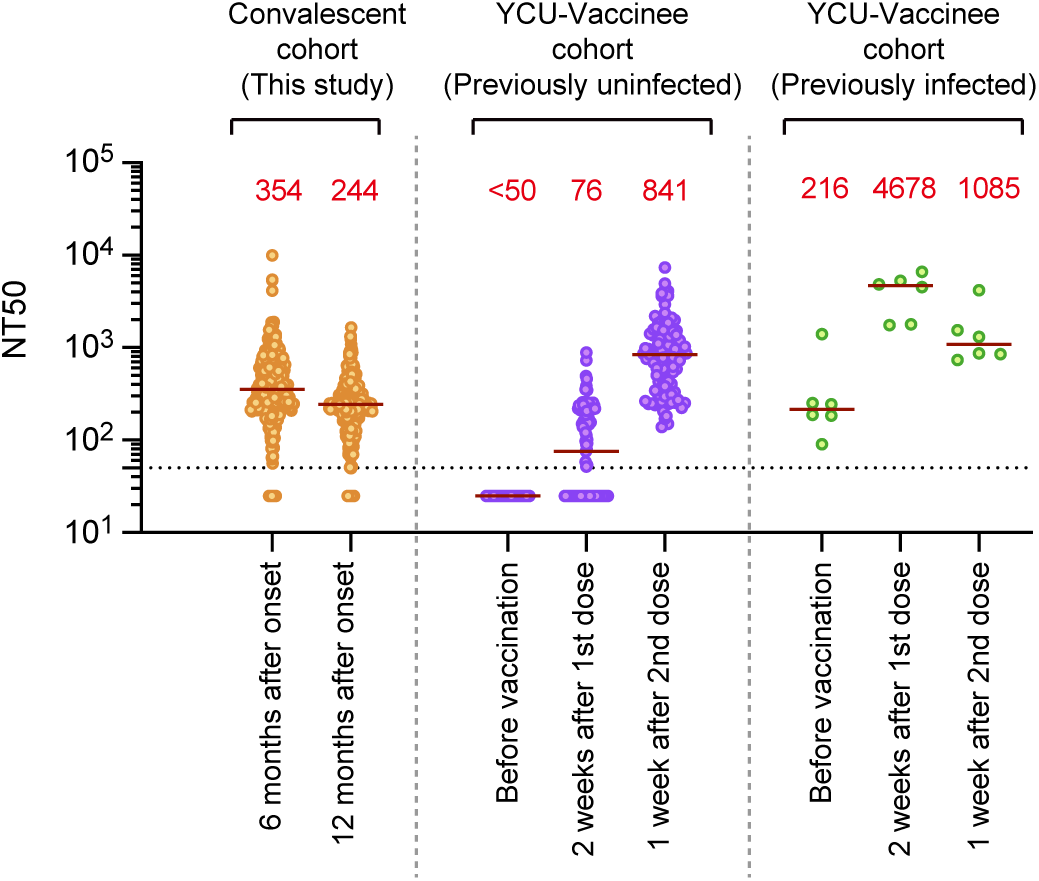
Comparison of neutralizing antibody titers between convalescent and vaccinee cohorts. Neutralizing antibody (NT50) of sera derived from the vaccine cohort was measured using the same method. Red letters in the graph indicate the median value of each antibody titer. Note that the YCU-vaccinee cohort (N=105 for previously uninfected, N=6 for previously infected) mainly consisted of healthcare workers, with a mean age of 42 years (ranging 24–62 years, 24% male).

**Supplementary Table S1.** Demographic and comorbidities during the acute phase.

|  | Overall<br>n=358 | Mild<br>n=264 | Moderate<br>n=69 | Severe<br>n=25 |
| --- | --- | --- | --- | --- |
| Age | 50.1 ± 13.2 | 47.4 ± 13.1 | 57.3 ± 10.3 | 59.4 ± 8.9 |
| Men | 195 (54.5%) | 123 (46.6%) | 51 (73.9%) | 21 (84.0%) |
| BMI | 23.8 ± 3.9 | 23.4 ± 3.9 | 24.5 ± 3.8 | 25.2 ± 3.2 |
| Never smoking | 216 (60.3%) | 174 (65.9%) | 30 (43.5%) | 12 (48.0%) |
| Fever | 305 (93.0%) | 214 (81.1%) | 66 (95.7%) | 25 (100%) |
| Cough | 156 (43.6%) | 113 (42.8%) | 35 (50.7%) | 8 (32.0%) |
| Dyspnea | 109 (30.4%) | 62 (23.5%) | 34 (49.31%) | 13 (52.0%) |
| Taste or smell disturbance | 177 (49.4%) | 150 (56.8%) | 22 (31.9%) | 5 (20.0%) |
| Diabetes | 28 (7.8%) | 10 (3.8%) | 14 (20.3%) | 4 (16.0%) |
| Hypertension | 62 (17.3%) | 32 (12.1%) | 21 (30.4%) | 9 (36.0%) |
| Cardiovascular disease | 17 (4.7%) | 9 (3.4%) | 7 (10.1%) | 1 (4.0%) |
| Cerebrovascular disease | 6 (1.7%) | 3 (1.1%) | 3 (4.3%) | 0 (0%) |

**Supplementary Table S2.** Authentic SARS-CoV-2 variants used in this study.

| Virus name | PANGO Lineage | GISAID Strain name | GISAID ID |
| --- | --- | --- | --- |
| WK-521 (WT) | A | hCoV-19/Japan/TY-WK-521/2020 | EPI_ISL_408667 |
| QHN001 (Alpha) | B.1.1.7 | hCoV-19/Japan/QHN001/2021 | EPI_ISL_804007 |
| TY8-612 (Beta) | B.1.351 | hCoV-19/Japan/TY8-612/2021 | EPI_ISL_1123289 |
| TY7-503 (Gamma) | P.1 | hCoV-19/Japan/TY7-503/2021 | EPI_ISL_877769 |
| TY11-927 (Delta) | B.1.617.2 | hCoV-19/Japan/TY11-927-P1/2021 | EPI_ISL_2158617 |
| TY11-330 (Kappa) | B.1.617.1 | hCoV-19/Japan/TY11-330-P1/2021 | EPI_ISL_2158613 |

**Supplementary Table S3.** Multiple regression analysis.

| Variables | Beta | 95% CI | p value |
| --- | --- | --- | --- |
| Age | 0.004 | 0.002, 0.007 | 0.002 |
| Severity |  |  | <0.001 |
| Mild | - |  |  |
| Moderate | 0.066 | -0.024, 0.155 | 0.2 |
| Severe | 0.283 | 0.148, 0.418 | <0.001 |
| Sex |  |  |  |
| Male | - |  |  |
| Female | -0.009 | -0.083, 0.065 | 0.818 |
| BMI | 0.006 |  | 0.233 |
| Smoking |  |  |  |
| Never | - |  |  |
| Former | -0.098 | -0.211, 0.014 | 0.086 |
| Current | -0.018 | -0.095, 0.059 | 0.645 |

## Supplementary Appendix

### Collaborating medical associations

*Tokyo Medical Association*

*Kanagawa Prefecture Medical Association*

*Osaka Medical Association*

### Collaborating doctors (ordered alphabetically by clinic affiliation)

*Dr. Isao Hayashi (Hayashi Clinic)*

*Dr. Koichi Hirahata (Hirahata Clinic)*

*Dr. Kiyomitsu Ikeoka (Ikeoka Clinic)*

*Dr. Hiroshi Kameoka (Kameoka Clinic)*

*Dr. Kazuo Suzuki (Kenkokan Suzuki Clinic)*

*Dr. So Ishii (Kudanshita Ekimae CoCo Clinic)*

*Dr. Ikuro Matsuba (Matsuba Medical Clinic)*

*Dr. Yoko Matsuzawa (Matsuzawa Diabetes Clinic)*

*Dr. Otoya Miho (Medical Corporation Yuhokai Miho Clinic)*

*Dr. Tetsuo Nishikawa (Nishikawa Clinic)*

*Dr. Kohei Azuma (Tsuzuki Azuma Clinic: Primary care and Rheumatology)*

*Dr. Koichiro Yamasaki (Yamasaki Family Clinic)*

*Dr. Yoshiaki Ono (Yokohama Respiratory Clinic)*

## Notes

### Author Declarations

All participants provided written informed consent and the study was approved by the Institutional Review Board of Yokohama City University (Reference No. B200600115, B200700023, B210300001).

## References

1. Polack FP, Thomas SJ, Kitchin N, et al. Safety and Efficacy of the BNT162b2 mRNA Covid-19 Vaccine. N Engl J Med 2020; 383:2603–15.

2. Galloway SE, Paul P, MacCannell DR, et al. Emergence of SARS-CoV-2 B.1.1.7 Lineage - United States, December 29, 2020-January 12, 2021. MMWR Morb Mortal Wkly Rep 2021; 70:95–9.

3. Tegally H, Wilkinson E, Giovanetti M, et al. Detection of a SARS-CoV-2 variant of concern in South Africa. Nature 2021; 592:438–43.

4. Fujino T, Nomoto H, Kutsuna S, et al. Novel SARS-CoV-2 Variant in Travelers from Brazil to Japan. Emerg Infect Dis 2021; 27.

5. Luchsinger LL, Ransegnola BP, Jin DK, et al. Serological Assays Estimate Highly Variable SARS-CoV-2 Neutralizing Antibody Activity in Recovered COVID-19 Patients. J Clin Microbiol 2020; 58.

6. Seow J, Graham C, Merrick B, et al. Longitudinal observation and decline of neutralizing antibody responses in the three months following SARS-CoV-2 infection in humans. Nat Microbiol 2020; 5:1598–607.

7. Wajnberg A, Amanat F, Firpo A, et al. Robust neutralizing antibodies to SARS-CoV-2 infection persist for months. Science 2020; 370:1227–30.

8. Characterisation WHOWGotC, Management of C-i. A minimal common outcome measure set for COVID-19 clinical research. Lancet Infect Dis 2020; 20:e192–e7.

9. Kubo S, Ohtake N, Miyakawa K, et al. Development of an Automated Chemiluminescence Assay System for Quantitative Measurement of Multiple Anti-SARS-CoV-2 Antibodies. Front Microbiol 2020; 11:628281.

10. Schmidt F, Weisblum Y, Muecksch F, et al. Measuring SARS-CoV-2 neutralizing antibody activity using pseudotyped and chimeric viruses. J Exp Med 2020; 217.

11. Gaebler C, Wang Z, Lorenzi JCC, et al. Evolution of antibody immunity to SARS-CoV-2. Nature 2021; 591:639–44.

12. Williamson EJ, Walker AJ, Bhaskaran K, et al. Factors associated with COVID-19-related death using OpenSAFELY. Nature 2020; 584:430–6.

13. Chia WN, Zhu F, Ong SWX, et al. Dynamics of SARS-CoV-2 neutralising antibody responses and duration of immunity: a longitudinal study. Lancet Microbe 2021; 2:e240–e9.

14. Greaney AJ, Loes AN, Crawford KHD, et al. Comprehensive mapping of mutations in the SARS-CoV-2 receptor-binding domain that affect recognition by polyclonal human plasma antibodies. Cell Host Microbe 2021; 29:463–76 e6.

15. Harvey WT, Carabelli AM, Jackson B, et al. SARS-CoV-2 variants, spike mutations and immune escape. Nat Rev Microbiol 2021; 19:409–24.

16. Li Q, Wu J, Nie J, et al. The Impact of Mutations in SARS-CoV-2 Spike on Viral Infectivity and Antigenicity. Cell 2020; 182:1284–94 e9.

17. Moriyama S, Adachi Y, Sato T, et al. Temporal maturation of neutralizing antibodies in COVID-19 convalescent individuals improves potency and breadth to circulating SARS-CoV-2 variants. Immunity 2021.

18. Miyakawa K, Jeremiah SS, Ohtake N, et al. Rapid quantitative screening assay for SARS-CoV-2 neutralizing antibodies using HiBiT-tagged virus-like particles. J Mol Cell Biol 2020; 12:987–90.

19. Miyakawa K, Stanleyraj JS, Kato H, et al. Rapid detection of neutralizing antibodies to SARS-CoV-2 variants in post-vaccination sera. J Mol Cell Biol 2021.

20. Betton M, Livrozet M, Planas D, et al. Sera neutralizing activities against SARS-CoV-2 and multiple variants six month after hospitalization for COVID-19. Clin Infect Dis 2021.

21. Goto A, Go H, Miyakawa K, et al. Sustained Neutralizing Antibodies 6 Months Following Infection in 376 Japanese COVID-19 Survivors. Front Microbiol 2021; 12:661187.

22. L’Huillier AG, Meyer B, Andrey DO, et al. Antibody persistence in the first 6 months following SARS-CoV-2 infection among hospital workers: a prospective longitudinal study. Clin Microbiol Infect 2021.

23. Yao L, Wang GL, Shen Y, et al. Persistence of Antibody and Cellular Immune Responses in COVID-19 patients over Nine Months after Infection. J Infect Dis 2021.

24. Wang Z, Muecksch F, Schaefer-Babajew D, et al. Naturally enhanced neutralizing breadth against SARS-CoV-2 one year after infection. Nature 2021; 595:426–31.

25. Long QX, Tang XJ, Shi QL, et al. Clinical and immunological assessment of asymptomatic SARS-CoV-2 infections. Nat Med 2020; 26:1200–4.

26. To KK, Tsang OT, Leung WS, et al. Temporal profiles of viral load in posterior oropharyngeal saliva samples and serum antibody responses during infection by SARS-CoV-2: an observational cohort study. Lancet Infect Dis 2020; 20:565–74.

27. Chen X, Pan Z, Yue S, et al. Disease severity dictates SARS-CoV-2-specific neutralizing antibody responses in COVID-19. Signal Transduct Target Ther 2020; 5:180.

28. Muller L, Andree M, Moskorz W, et al. Age-dependent immune response to the Biontech/Pfizer BNT162b2 COVID-19 vaccination. Clin Infect Dis 2021.

29. Frieman M, Harris AD, Herati RS, et al. SARS-CoV-2 vaccines for all but a single dose for COVID-19 survivors. EBioMedicine 2021; 68:103401.

30. Muecksch F, Weisblum Y, Barnes CO, et al. Affinity maturation of SARS-CoV-2 neutralizing antibodies confers potency, breadth, and resilience to viral escape mutations. Immunity 2021.

